# Thirteen-fold variation between states in clozapine prescriptions to United States Medicaid patients

**DOI:** 10.1101/2022.04.03.22273352

**Authors:** Rizelyn A. Benito, Michael H. Gatusky, Mariah W. Panoussi, Kenneth L. McCall, Anisa S. Suparmanian, Brian J. Piper

## Abstract

**Background:** Clozapine was the first atypical antipsychotic for treating schizophrenia, with a long history of controversy over its usage. Guidelines currently recommend clozapine for patients diagnosed with refractory schizophrenia. However, this agent may be underutilized because of the costs associated with close monitoring of its adverse effects, particularly agranulocytosis. This is unfortunate because clozapine has demonstrated greater effectiveness compared with other antipsychotics. It is essential to examine clozapine usage to determine if it is being adequately utilized among United States (US) Medicaid patients.

**Methods:** Medicaid data, including the number of quarterly clozapine prescriptions and the number of Medicaid enrollees in each state from 2015-2019, was collected and used to evaluate clozapine use over time. Data-analysis and figures were prepared with Excel and GraphPad Prism. Exploratory correlations were completed between prescriptions per enrollee and other factors.

**Results:** The number of prescriptions, corrected for the number of enrollees in Medicaid, was generally consistent over time. However, average prescriptions per quarter were markedly lower in 2017 compared with other years, decreasing by 44.4% from 2016 average prescriptions per quarter. From 2015 to 2019, states from the upper Midwest and Northeast regions of the country had the highest average clozapine prescriptions per 10,000 Medicaid enrollees (ND: 190.0, SD: 176.6, CT: 166.2). States from the Southeast and Southwest had much lower average rates (NV: 17.9, KY: 19.3, MS: 19.7). There was an over ten-fold difference in clozapine prescriptions between states from 2015-2019 (2015 = 19.9-fold, 2016=11.4 fold, 2017=11.6 fold, 2018=13.3 fold, and 2019=13.0 fold). There was a moderate correlation of (r(48) = 0.49, *p* < .05) between prescriptions per 10,000 enrollees and the Medicaid spending per enrollee in each state in 2019. There was a small, but significant, correlation between prescriptions per enrollee and percent white (r(48) = 0.30, *p* < .05).

**Conclusion:** Clozapine is an important pharmacotherapy for refractory schizophrenia. Overall, clozapine use tends to be highest among the upper Midwest and Northeast states. Further research is ongoing to better understand the origins of the thirteen-fold regional disparities in clozapine use in 2019 and the state level variation in Medicaid spending.

## Introduction

Clozapine is a second-generation antipsychotic (SGA) and is also generally known as an atypical antipsychotic (1). Current clinical guidelines state that clozapine is an effective treatment for those with treatment-resistant schizophrenia (TRS) (2). Presently, the clinical and research criteria are failure of two trials of non-clozapine antipsychotics of standard dose and duration (3). However, current evidence demonstrates a lack of clozapine utilization by providers (4). Clozapine has been known to cause numerous adverse effects that range from hyperglycemia and weight gain to more serious reactions such as seizures and myocarditis (1). One particular adverse effect correlates with drug induced agranulocytosis in patients which leads to increased susceptibility and death from infectious diseases (5).

Another factor that further complicates this controversy over clozapine is the high cost associated with close monitoring for agranulocytosis (2). To put this into perspective, the financial burden schizophrenia imposes annually on patients is approximately $23 billion (6). Direct healthcare costs in the United States (U.S.) are estimated to be 3 to 11-fold higher for TRS patients, which includes multiple hospitalizations (3). Nevertheless, recent research has revealed clozapine along with other SGAs as more cost effective (7).

Besides adverse effects, other impediments influencing clozapine underutilization include complications when administration and registry changed from individual pharmaceutical companies to a single clozapine program (8). The new single registry has new requirements and procedures that have hindered patient initiation on clozapine (8). For example, it has interfered with provider collaborations specifically when transitioning a patient’s care (8). Another aspect is inadequate centralized resources to benefit patients and their families with services such as patient education and adherence monitoring (8). Additionally, failure to account for benign ethnic neutropenia when considering clozapine for African-American patients as well as the scarcity of adequate psychiatric services in correctional facilities are obstacles leading to clozapine underutilization (8).

There’s also the interference of physician’s knowledge, perspectives, and attitudes affecting clozapine usage in various countries (9-10). Recent surveys have shown that 40.5% and 64.7% of physicians prefer other antipsychotics or combine two antipsychotics before considering clozapine (9,11). One survey discovered that 66.0% of psychiatrists stated that their patients were less satisfied when treated with clozapine compared to those treated with other atypical antipsychotics (11). Specifically in the US, researchers had 295 providers complete a questionnaire where 33.0% of providers reported prescribing clozapine after three or more unsuccessful antipsychotics have been tried (12). There was also unanimous physician reluctance with prescribing clozapine because of inadequate knowledge or experience with clozapine and concerns with patient compliance (8–10,12).

The barriers described above are causing a visible variation in clozapine use throughout the US. One study noted that 4.8% of schizophrenic patients were on clozapine with a slight decline during 2001-2005 (13). This analysis revealed that clozapine was used least by Deep South states, while states in New England, the Rocky Mountain region, and Washington had comparatively frequent use (13). A more recent report in 2016 confirmed this previous data and determined that 15.6% of Medicaid recipients with schizophrenia in South Dakota received clozapine compared to 2.0% in Louisiana (13,14).

Therefore, there is a need for an updated examination of the variation of clozapine use in the US. Past studies have endeavored to uncover possible reasons for this variation, yet it still remains ambiguous (13,14). Thus, the aim of our research was to conduct a secondary analysis of Medicaid data from 2015-2019 to further investigate the underlying cause(s) of clozapine usage variation throughout the US.

## Methods

### Procedures

The data was obtained from the Data.Medicaid.gov database for 2015 to 2020 (15), which focused on the nationwide drug use of clozapine. Data was collected for clozapine and the brand name counterparts (Versacloz, Clozaril, and Fazaclo). All obtained data was next placed into Excel spreadsheets to organize and proceed with data-analysis. Procedures were approved by the IRB of the University of New England and Geisinger.

### Data-analysis

The number of prescriptions per 10,000 Medicaid enrollees was calculated to find a standardized prescription rate for each state using Excel. Bar graphs and heat maps were used to analyze the overall trends of clozapine use by each state. Line graphs were used to visualize clozapine prescription rates in each quarter from 2015-2020. The 2019 Medicaid data was used to calculate the dollars spent per Medicaid enrollee, and the Pearson correlation was subsequently determined between dollars spent per enrollee and clozapine prescription rates. GraphPad Prism was used to create these figures.

The 95% confidence intervals were also calculated for the prescriptions per 10,000 enrollees. The fold difference was calculated between the states with the highest and lowest average prescriptions per 10,000 from 2015-2019. Pearson correlation coefficients were calculated for amount spent per enrollee, percent white population, and percent rural population in each state compared with prescription rates.

## Results

Clozapine prescriptions remained relatively constant from 2015 through the first half of 2020 with a considerable drop in total Medicaid prescriptions in 2017. Figure 1 shows the overall number of clozapine prescriptions per quarter during this timeframe, including a 44.4% drop from 2016 average prescriptions per quarter to 2017 prescriptions per quarter. The highest amount of clozapine being prescribed was during the third quarter of 2016, with 173,087 prescriptions. The lowest amount of clozapine prescribed was during the third quarter of 2017, with 80,070 prescriptions.

**Figure 1.**
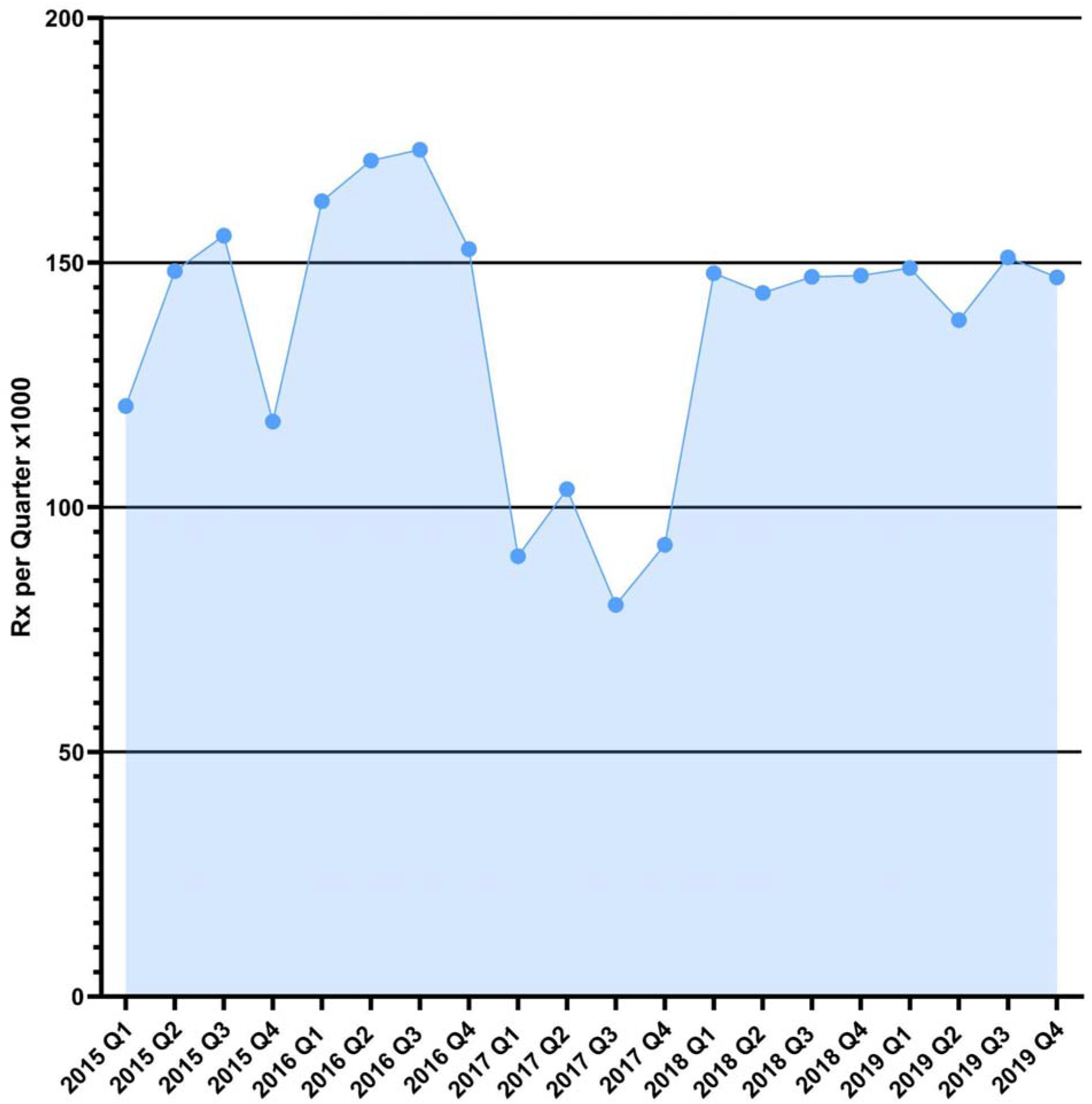
Number of Medicaid prescriptions per quarter for clozapine during 2015 to quarter 2 of 2020.

We also found a stark difference between generic and brand name clozapine prescriptions with the vast majority of clozapine prescriptions being generic. In 2019 579,875 of 585,217 prescriptions or 99.09% of nationwide Medicaid prescriptions were generic.

Upon calculating the prescriptions of clozapine in each state per 10,000 Medicaid enrollees, there were substantial differences among states’ prescription rates. Figure 1 shows this nationwide comparison for 2019. South Dakota had the highest rate of prescribing clozapine (191.6). The lowest prescribing state was Arkansas (14.7). The average rate was 79.3 prescriptions per 10,000 Medicaid enrollees with a standard deviation of 43.7. The 95% confidence interval was from -6.4 to 165.0 prescriptions per 10,000 enrollees with North Dakota, South Dakota, and Missouri being significantly elevated. Finally, there was a 13.0 fold difference between the lowest and highest prescribing states, Arkansas and South Dakota respectively.

Results were similar when averaged from 2015-2019. Overall, North Dakota had the highest rate (190). The lowest prescribing state was Nevada (17.9). The average rate was 80.4 with a standard deviation of 44.7. The 95% confidence interval was from - 7.2 to 168 with North and South Dakota above the range of the confidence interval. Finally, there was a 10.6-fold difference between the lowest and highest prescribing states, Nevada and North Dakota respectively.

Figure 2A and 2B represents usage of clozapine per 10,000 Medicaid enrollees per state in 2018 and 2019 respectively and depicts highest prescriptions in the midwest and western states. As shown in Figure 2A, in 2018 North Dakota had the highest clozapine prescription rate (221.9) while the lowest rate was Nevada (16.7). Figure 2B shows that South Dakota (191.6) had the highest and Arkansas had the lowest (14.8) clozapine prescription rate in 2019.

**Figure 2.**
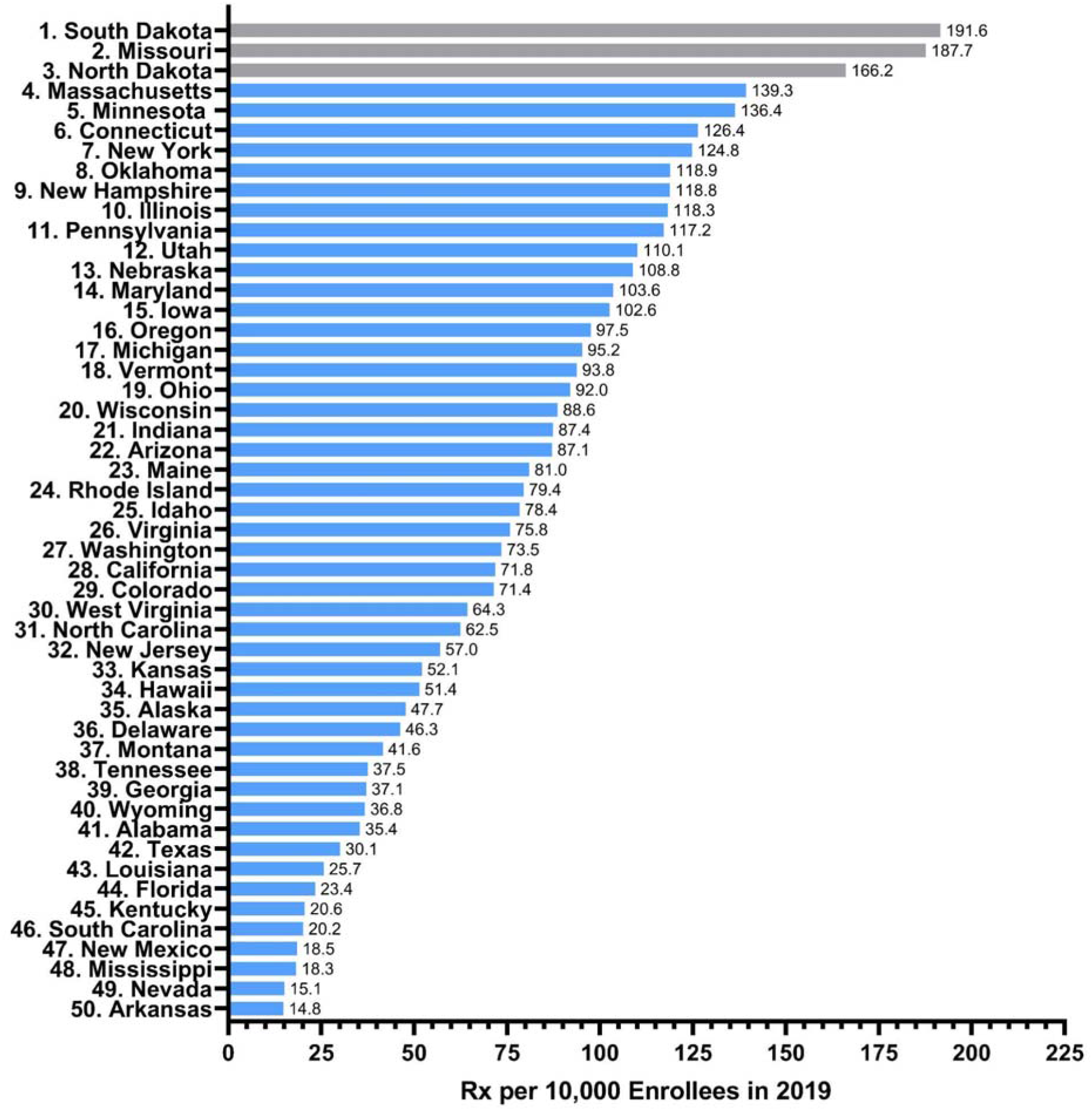
State use of clozapine, ranked, per 10,000 Medicaid enrollees for 2019. Gray states were significantly elevated (p < .05) relative to the national mean (79.3 prescriptions with a standard deviation of 43.7, 95% Confidence interval = -6.4 to 165.0).

Additionally, we found a significant correlation between Medicaid spending and the clozapine prescription rates of states. Figure 4 demonstrates this positive correlation between clozapine prescriptions per 10,000 enrollees and the Medicaid spending per enrollee in each state in 2019 (r(48)= +0.49, *p* < .05).

**Figure 3.**
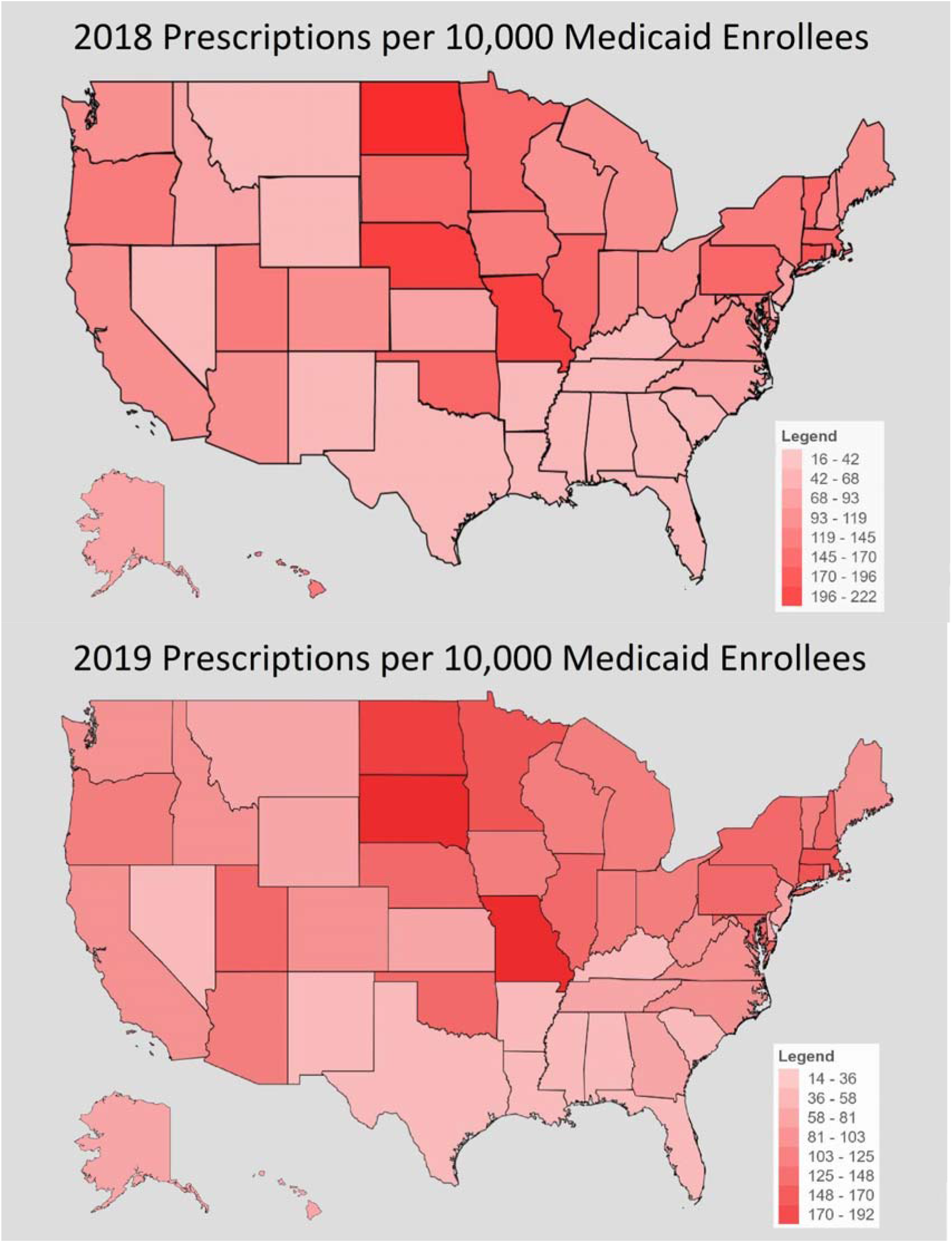
Heatmap of clozapine usage per 10,000 Medicaid enrollees for 2018 (A) and 2019 (B).

**Figure 4.**
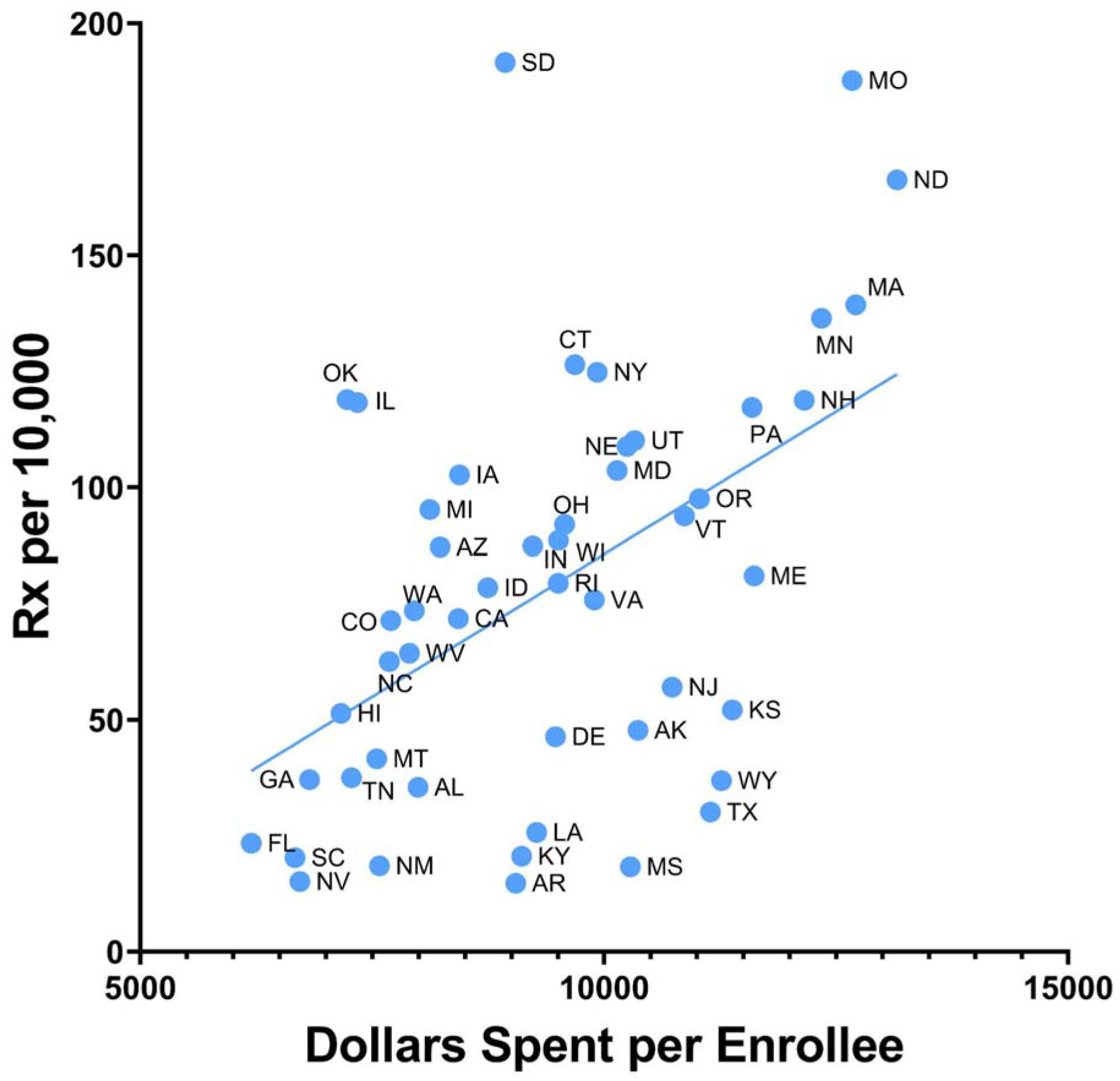
Scatterplot between clozapine prescriptions per 10,000 enrollees and the Medicaid spending per enrollee in each state in 2019 (r(48) = +0.49, *p* < .05).

Pearson coefficients were also calculated between states’ rural population percentage according to the 2010 Census and clozapine prescriptions per 10,000 Medicaid enrollees. The correlations were minimal at 0.072, 0.071, -0.004, -0.034, and - 0.021 from 2015 through 2019, respectively (not shown). Small positive Pearson coefficients were calculated between states’ percent white population and clozapine prescriptions per 10,000. Figure 5 illustrates this correlation in 2019 with a correlation of 0.38 (p < .05). The coefficients for 2015-2018 were 0.36, 0.44, 0.34, and 0.32 (all p < .05) respectively.

**Figure 5.**
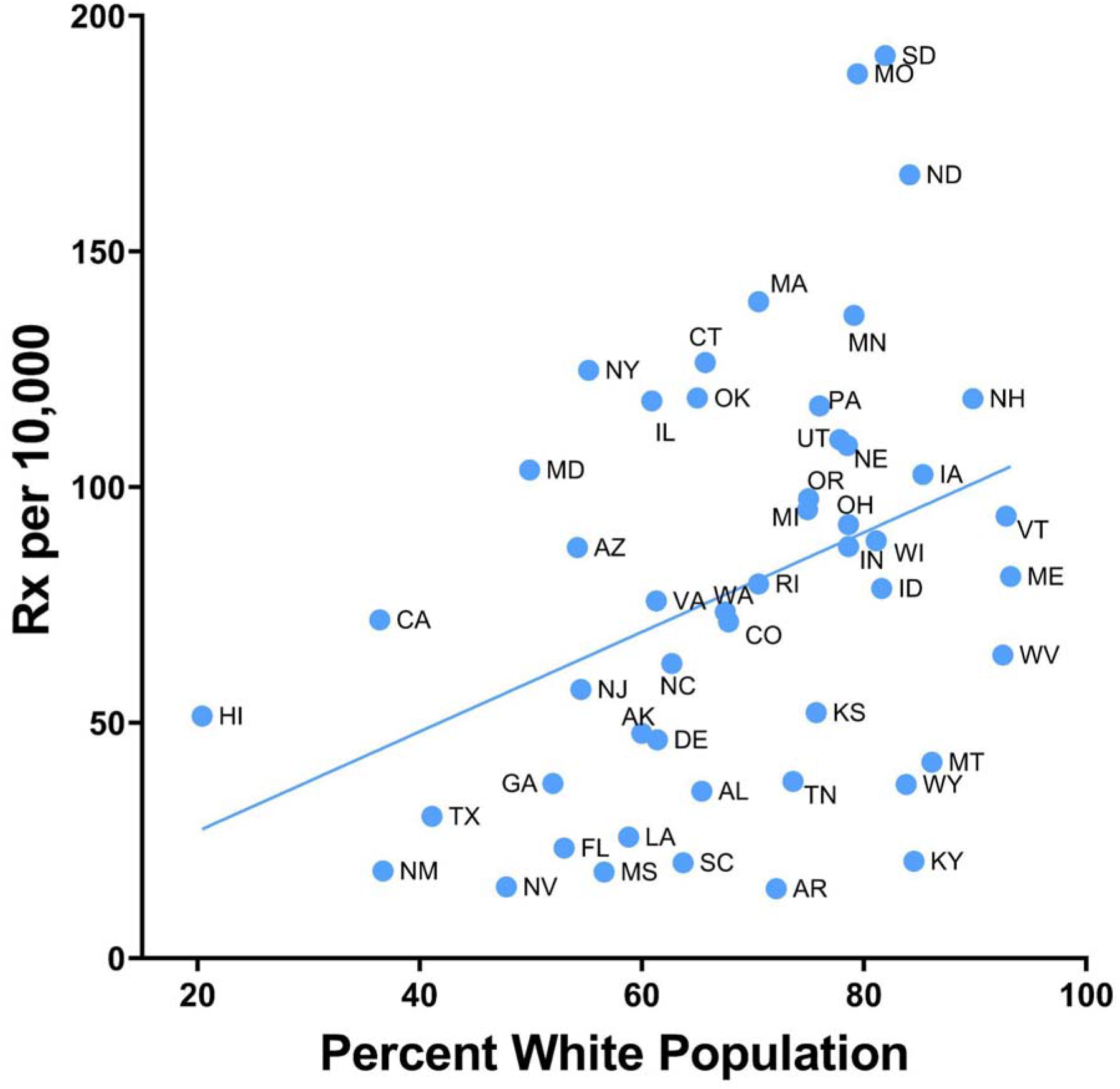
Scatterplot between clozapine prescriptions per 10,000 enrollees and the Medicaid spending per enrollee in each state in 2019 (r(48) = +0.38, *p* < .05).

## Discussion

There are several key findings to this examination of clozapine prescribing to US Medicaid patients. This study identified pronounced (ten+ fold) regional disparities in clozapine prescriptions to Medicaid patients from 2015 to 2019. States in the Southeast and Southwest US had lower clozapine prescription rates with Nevada, Kentucky, Mississippi, New Mexico, and Florida having the lowest average rates. Past studies cited rural geographic location as an influence on both the prevalence of TRS and on the availability of psychiatric treatment (3,4,16). Investigators found that living in a rural region was a predictor for TRS, suggesting that states with higher rural populations may have higher clozapine prescription rates (3). Conversely, the Health Resources and Services Administration reported that rurality was the most common marker of US counties with primary care health professional shortages (16). This suggests that counties with higher rural populations may also lack access to psychiatric care and thus receive less clozapine prescriptions proportionally. However, our analysis showed no significant correlation between percent rural population per state and clozapine prescriptions per state.

Several low prescription states in the Southeast and Southwest tended to have higher non-white populations compared with higher prescription rate states such as North and South Dakota. Of the five states with the lowest prescription rates, all but Kentucky were in the top third of states with the highest non-white populations in the US according to Kaiser Family Foundation estimates for 2019 (17). Conversely, North and South Dakota were in the bottom third for non-white population. Additionally, our analysis revealed small to moderate (0.32 to 0.48) significant positive correlations in all years between percent white population in a state and clozapine prescription rates. Overall, the ten-fold variation among the states in clozapine prescriptions aligns with past data showing that being white was a factor significantly associated with clozapine initiation (18).

Furthermore, states in the Southeast have among the highest Black populations in the US. There is also data showing that Black Americans were less likely to be prescribed clozapine than White Americans (19). In addition to factors such as prescriber bias and the anticipation of nonadherence, the presence of benign ethnic neutropenia may impact prescription rates among Black patients (19, 20). Benign ethnic neutropenia is an unexplained neutrophil count of < 1.5 × 10^9^/L, which does not confer an increased risk of infection and is commonly seen in individuals of African and Afro-Caribbean descent, as well as some Middle Eastern ethnic groups (8,19). Although the US’s threshold for clozapine discontinuation is lower than that of the United Kingdom and FDA monitoring criteria were revised in 2015, there is still evidence that clozapine is under-prescribed in Black populations and that Black patients are more likely to discontinue clozapine (8,19, 20). Practitioners may have been slow to adopt these new recommendations due to a lack of product labeling and concerns about using lower neutrophil count thresholds (8). Furthermore, benign ethnic neutropenia is a diagnosis of exclusion and is likely underdiagnosed (19).

This data analysis led to speculation that Medicaid expansion during the early 2010s would be linked with the upward trend in clozapine usage. Surprisingly, there was a sizable decrease in clozapine prescriptions during 2017. One possible explanation for this sudden d ecrease could be related to the addition of long-acting injectable (LAI) antipsychotics to Medicaid’s preferred list in some states. For example, the Pennsylvania Department of Human Services Preferred Drug List now has haloperidol lactate syringe as a preferred agent (21). LAIs improve patient adherence and reduce healthcare costs (22–25). Three of these four studies used Medicaid data to review the effectiveness of LAIs and cost, and each concluded that these drugs improved patient adherence and overall costs were reduced due to decreased hospitalizations (23–25). Thus, LAIs are being seen as more beneficial due to their efficiency in managing schizophrenia (22, 24). Interestingly, one research group stated that oral antipsychotics were similar in cost effectiveness to LAIs, while another found LAIs to reduce healthcare costs by half when compared to oral antipsychotics (23, 25). Hence, further investigation is required to fully determine if there is a difference between LAIs and SGA oral antipsychotic’s efficiency in managing TRS and schizophrenia along with their related healthcare costs.

There is a need to carefully consider and potentially overcome the factors that contribute to variation in clozapine usage. One solution could be to increase the involvement of advanced practitioners and primary care physicians in the close monitoring of patients on clozapine. Shared care with primary care physicians in the close monitoring of clozapine’s side effects can effectively minimize the likelihood of agranulocytosis (26). Efficient communication among a psychiatrist, primary care physician, advanced practitioner, and pharmacist is also imperative to effectively monitor clozapine’s side effects (26, 27). In terms of safety and efficiency, delegating clozapine monitoring to advanced practitioners has been deemed to be equivalent to a physician monitoring clozapine (28, 29). Therefore, increased involvement in primary care and advanced practitioners may increase the usage of clozapine among specialists.

Another strategy to increase clozapine usage is to improve education on the advantages of clozapine and the ability to safely monitor its side effects. In one survey of psychiatrists, trainees, and advanced practitioners completed in September 2019, 68.2% of respondents viewed clozapine usage as a burden (30). In the former survey along with another that surveyed physicians vs nurse practitioners, both found that there was little to no difference of physicians and advanced practitioners viewpoints in regards to clozapine under-prescription (30, 31). Thus, improved education to current and future providers on the value of prescribing clozapine along with involving primary care and advanced practitioners may be the key to improving clozapine usage.

Lastly, a few more solutions would be to involve outpatient services and the development of a point of care monitoring device. One study discovered that areas with high utilization of clozapine were related to the integration of non-psychiatric providers, organized mental health management, and clozapine clinics (32). On the other hand, areas with low clozapine utilization had a lack of organized outpatient management and clinics as well as limited care facilities (32). Another benefit of organized outpatient clinics is that it reduces the number and cost of hospitalizations by more than half (33). Finally, there needs to be a point of care device that provides patients the ability to check clozapine levels and white blood cell counts at home (34). This device would significantly improve prescriber utilization as well as patient adherence and willingness to use clozapine (34). Hence, improved clozapine usage among prescribers must include the involvement of primary care and advanced practitioners, improved education on the benefits of clozapine, organized outpatient clinics for monitoring clozapine, and the development of a clozapine and white blood cell count point of care device.

Some caveats and future directions are noteworthy. A limitation of this study is that Medicaid data for 2020 was incomplete at the time of data collection and analysis. There is also limited knowledge regarding the specific reasons for the drop in clozapine usage during 2017. Further studies that include electronic medical records will be required to further characterize this change in the clozapine. Further updates on this research among Medicaid and Medicare patients are needed. Future directions of this research also include an evaluation of the trends in utilization of generic and brand name clozapine. This would be invaluable knowledge, with potential safety concerns generic clozapine as there have been cases of schizophrenia relapse or exacerbation when substituting generic for branded clozapine (35). It is also problematic because as shown in Figure 4, 99.1% of prescriptions in 2019 were for generic clozapine. Evidence has shown that when some patients are switched from Clozaril to a generic form, they exhibit relapse or exacerbation of schizophrenic symptoms (35, 36). Therefore, research in this area would clarify if generic clozapine is harmful and if so, could lead to safer and efficacious versions that are less costly. Additionally, it would be valuable to research the costs of medication between different types and states in the future. It may also be possible that costs and rates of reimbursement for clozapine prescriptions, as well as the lab work necessary to monitor possible agranulocytosis, may have influenced prescription rates across the US. For instance, psychiatrists in states with lower clozapine costs or higher rates of reimbursement for hematological monitoring may be more likely to prescribe it. Lastly, the effects of step therapy to clozapine use throughout the years may affect the rates of clozapine prescription and should be analyzed in future research. Step therapy requires that patients undergo a series of treatments (generally lower cost) prior to prescription of another drug such as clozapine (37). It is likely that clozapine prescription rates will differ significantly in states where clozapine prescription is restricted by step therapy.

## Conclusion

There was over a ten-fold difference between the state’s clozapine usage. There were lower average clozapine prescription rates in the Southeast and Southwest, which can be attributed to numerous factors. Significant associations with Medicaid spending and race were identified. Factors that are possibly influencing clozapine prescription numbers are physician reluctance, serious adverse effects (i.e. agranulocytosis), and LAIs added to Medicaid’s preferred drug list. A possible solution to increase clozapine usage via improving prescriber’s and patient’s ability to manage and adhere to clozapine monitoring. Therefore, future investigation is required to elaborate upon these influential factors as well as implementing the aforementioned solutions in order to decrease disparities in clozapine usage among the states.

## Data Availability

Raw data is available at: https://www.medicaid.gov/medicaid/prescription-drugs/state-drug-utilization-data/index.html

## Acknowledgements

We would like to thank the Biomedical Research Club for their support. BJP is supported by the Health Resources Services Administration (D34HP31025). The National Institute of Environmental Health Sciences (T32 ES007060-31A1) provided software used for figure construction.

## Supplemental Figures

**Supplemental Figure 1.**
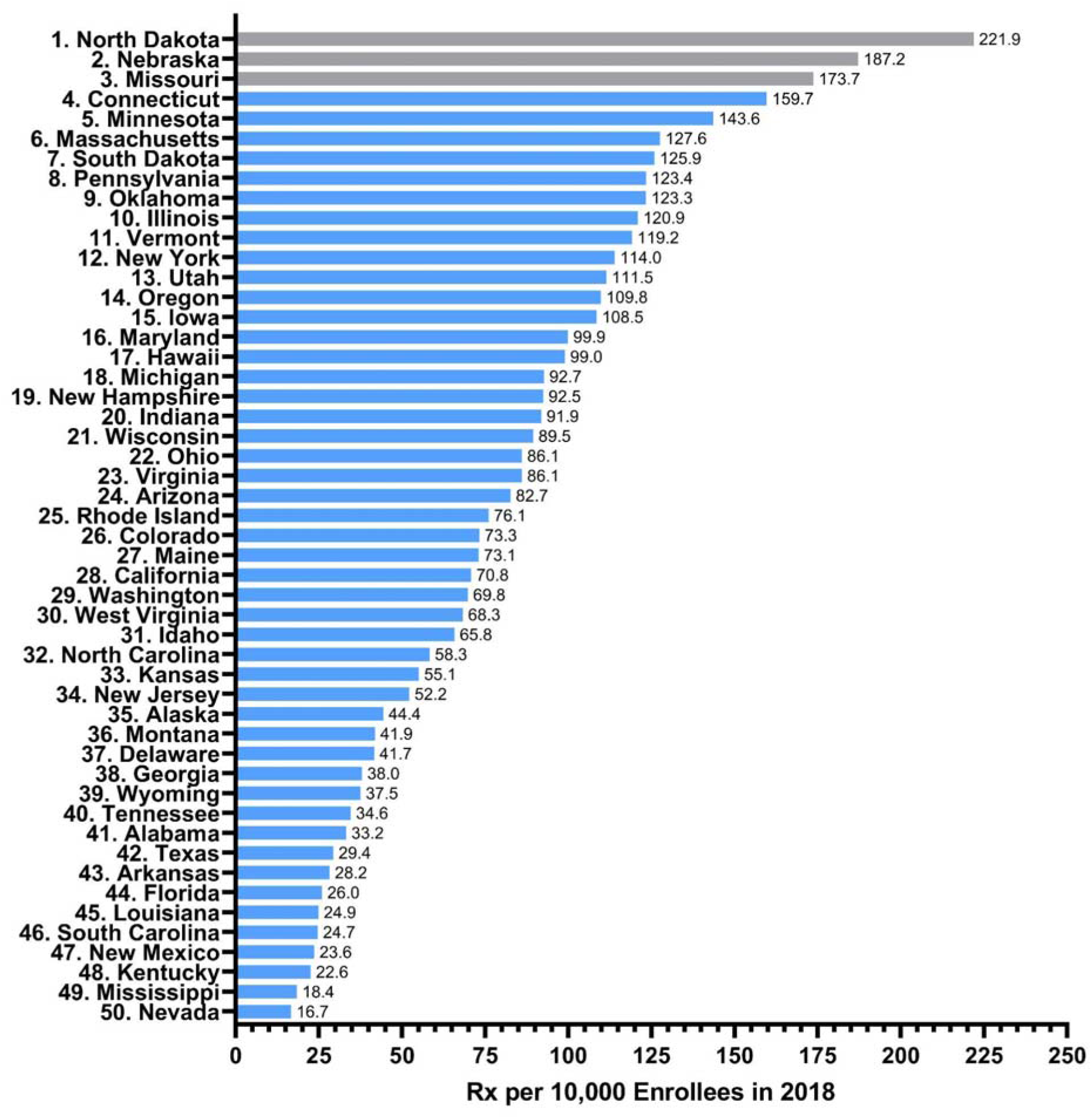
State use of clozapine, ranked, per 10,000 Medicaid enrollees for 2018. States in gray were significantly elevated (*p* < .05) relative to the national mean (82.1 prescriptions with a standard deviation of 46.6, 95% Confidence interval = - 9.2 to 173.3).

**Supplemental Figure 2.**
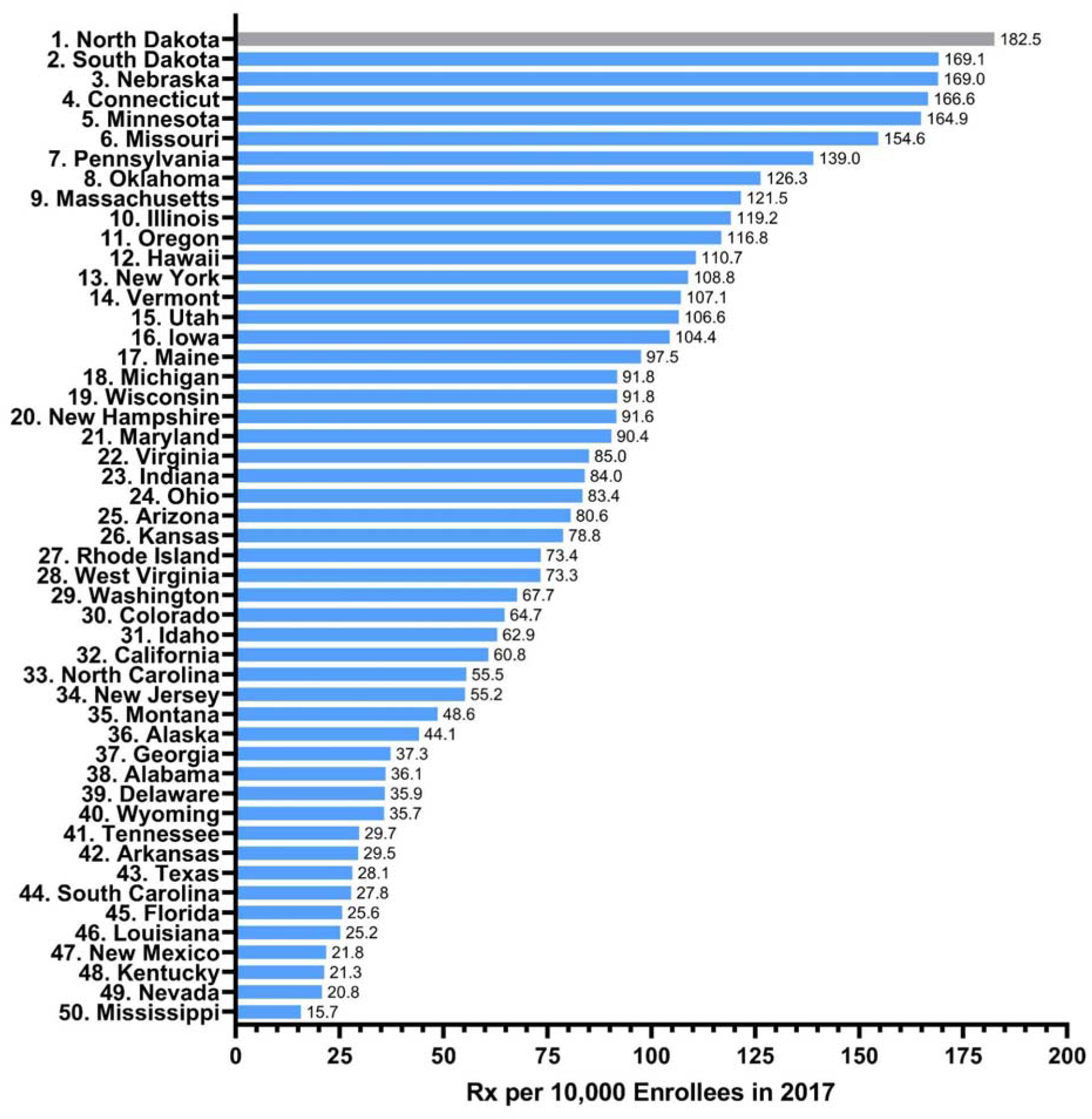
State use of clozapine, ranked, per 10,000 Medicaid enrollees for 2017. State in gray were significantly elevated (*p* < .05) relative to the national mean (82.1 prescriptions with a standard deviation of 45.3, 95% Confidence interval = -6.6 to 170.8).

**Supplemental Figure 3.**
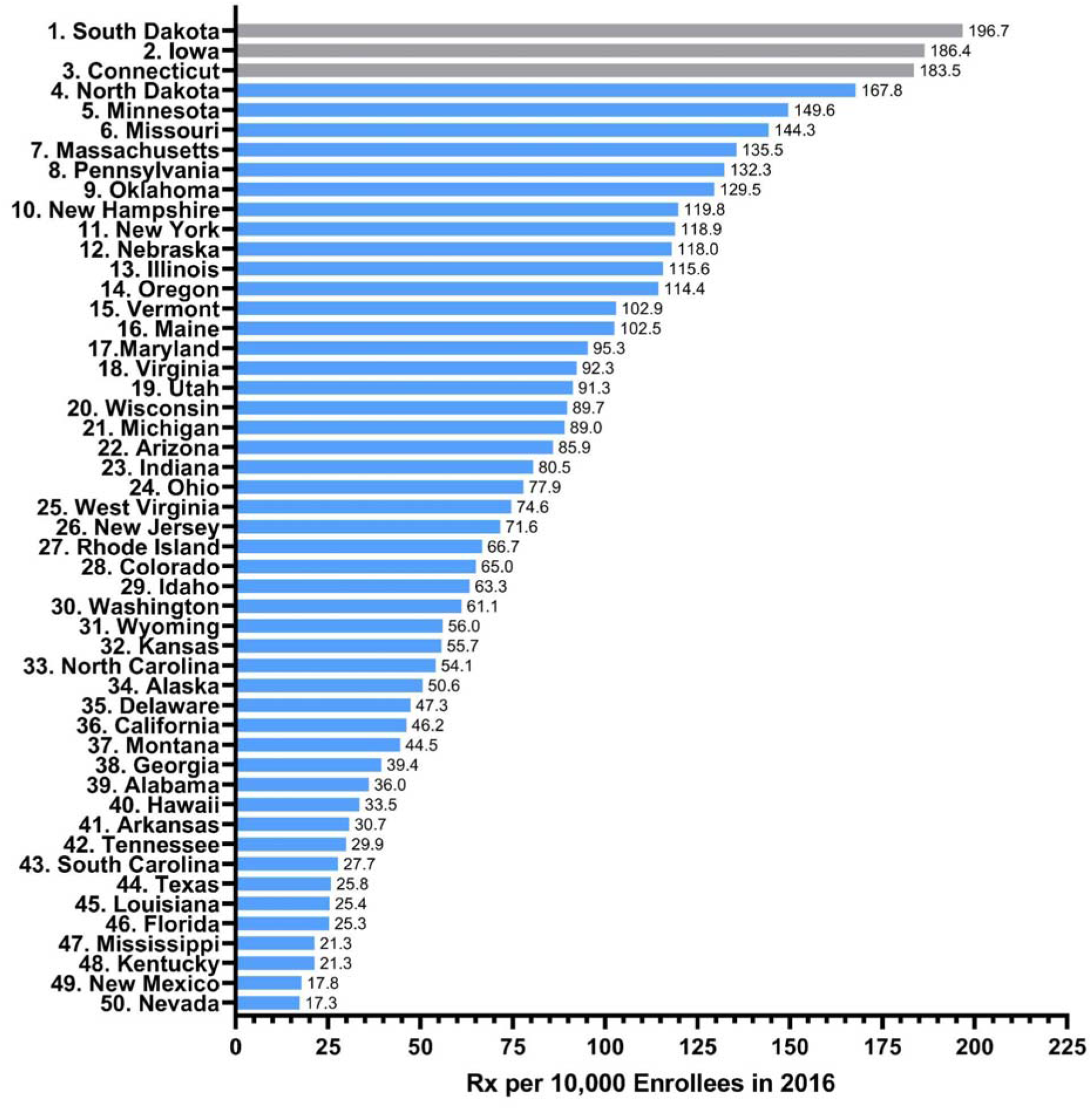
State use of clozapine, ranked, per 10,000 Medicaid enrollees for 2016. States in gray were significantly elevated (*p* < .05) relative to the national mean (81.8 prescriptions with a standard deviation of 47.3, 95% Confidence interval = - 10.9 to 174.5).

**Supplemental Figure 4.**
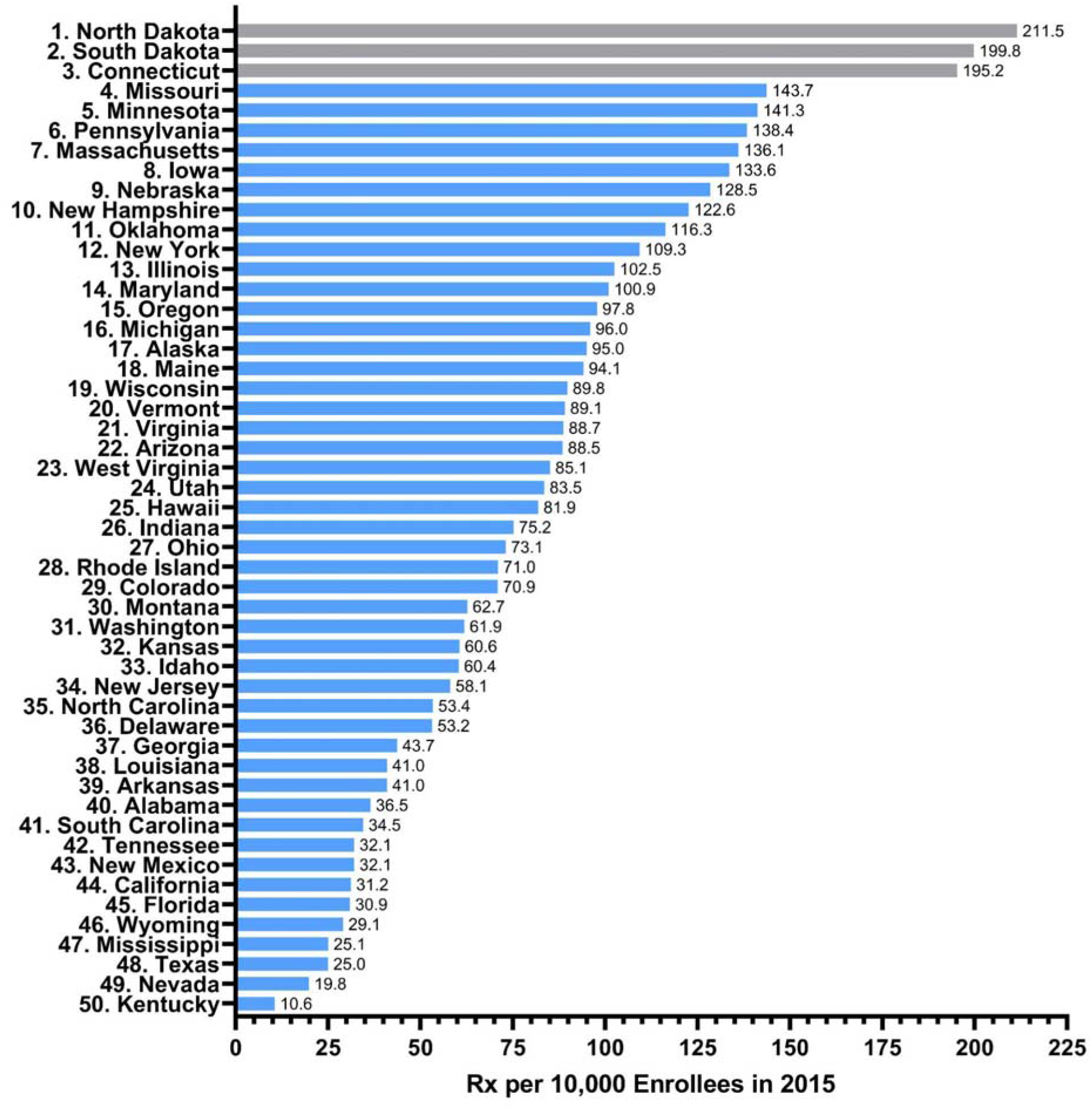
State use of clozapine, ranked, per 10,000 Medicaid enrollees for 2015. States in gray were significantly elevated (*p* < .05) relative to the national mean (83.5 prescriptions with a standard deviation of 46.0, 95% Confidence interval = - 6.6 to 173.6).

